# Data-efficient Self-Supervised Diffusion Learning for Detecting Myofascial Pain in Upper Trapezius Muscle with B-mode Ultrasound Videos

**DOI:** 10.64898/2026.04.07.26350333

**Authors:** Hao-En Lu, David Koivisto, Yuyang Lou, Zixue Zeng, Tong Yu, Jing Wang, Xin Meng, Cameron Nowikow, Richard Wilson, Dinesh Kumbhare, Jiantao Pu

## Abstract

Deep learning has transformed medical image and video analysis, but it usually requires large, well annotated datasets. In many clinical domains, especially when testing novel mechanistic hypotheses, such retrospective datasets are hard to obtain since acquiring adequate cohorts is time intensive, costly, and operationally difficult. This creates a critical translational gap: scientifically compelling early stage ideas may remain untested due to lack of sufficient sample size to support conventional deep learning pipelines. Developing data-efficient strategies for evaluating new hypotheses within small prospective cohorts is therefore essential to de-risk innovation before large-scale validation. Myofascial Pain Syndrome (MPS) exemplifies this challenge, as quantitative ultrasound imaging biomarkers for MPS remain underexplored. We investigated whether MPS in the upper trapezius can be detected from full B-mode ultrasound videos in a small prospective cohort (11 controls, 13 patients). Videos were automatically preprocessed and resampled using a sliding window strategy to expand training samples (404 clips). A self-supervised Video Diffusion Encoder (VDE) is developed to learn spatiotemporal representations without relying on extensive labeled data, and compared it with transfer-learning-based ResNet, VideoMAE, and SimCLR. Using subject-level stratified four-fold cross-validation, the VDE outperformed transfer learning baselines and achieved performance comparable to SimCLR, with subject-level AUC of 0.79 (0.66–0.90) and accuracy of 0.86 (0.79–0.91), and no significant differences between latent-only and combined trigger point analyses. These results demonstrate that self-supervised diffusion learning can support robust, data-efficient deep learning in small prospective studies, enabling early feasibility testing of innovative ultrasound biomarkers before large-scale clinical trials.

## 1. Introduction

Myofascial Pain Syndrome (MPS) is a chronic musculoskeletal condition characterized by regional pain and commonly linked to myofascial trigger points (MTrPs), localized contraction nodules within taut muscle bands that cause spontaneous or stimulus-evoked pain. Accurate identification of MTrPs are essential for effective diagnosis of MPS (Dua and Chang 2025; Harden 2007). In clinical practice, manual palpation remains the most common method for assessing MTrPs, however, this approach is highly subjective (Lucas et al., 2009; Rathbone et al., 2017). Ultrasonography has gained attention as a promising tool that offers dynamic visualization of MTrPs and surrounding tissues. While some studies describe MTrPs as elliptical hypoechoic regions on B-mode ultrasound (Sikdar et al., 2009), these findings remain inconsistent. Prior studies have relied on handcrafted texture features (Behr et al., 2019; Shomal Zadeh et al., 2023) or convolution neural networks (Koh et al., 2023) for MTrPs identification using static ultrasound frames containing hypoechoic MTrPs. However, these approaches introduce subjectivity and risks omitting important temporal or dynamic information present across the full video. As such, it is desirable to use entire ultrasound videos to capture tissue structure characteristics associated with myofascial pain for better diagnosis.

Deep Learning (DL) models have achieved state-of-the-art performance in natural video classification (Arnab et al., 2021; Hara et al., 2017) and shown increasing propose in medical imaging applications (Shea et al., 2023). Nevertheless, DL typically require large, well-annotated datasets to achieve robust generalization, which poses a fundamental barrier when testing novel clinical ideas where retrospective datasets are unavailable and prospective studies are constrained by slow patient enrollment and the high cost of expert annotation. Thus, large-scale prospective datasets are often unavailable during the initial exploration of novel clinical applications, highlighting the need for data-efficient strategies that can operate reliably under limited-sample conditions.

To reduce reliance on labeled data, transfer learning and self-supervised learning (SSL) have been widely adopted. Transfer learning leverages pretraining on large source-domain datasets to improve generalization in smaller target-domain data (Jain et al., 2024; Kim et al., 2022), while SSL learns data representation through unlabeled surrogate tasks (Ericsson et al., 2022; Krishnan et al., 2022). Earlier SSL approaches focused on contrastive learning (Chen et al., 2020) and masked autoencoder (He et al., 2022; Tong et al., 2022), but recently, diffusion models (Ho et al., 2020) have emerged as effective alternatives for SSL. Studies have demonstrated that their multi-level denoising process can be leveraged to extract meaningful features for downstream tasks (Fuest et al., 2024; Han et al., 2024; Mukhopadhyay et al., 2024; Xiang et al., 2023). Notably, these methods are designed primarily to improve performance on moderately sized retrospective datasets, while few studies focus on developing frameworks for evaluating early-stage ideas with small video datasets.

In this study, we investigate the feasibility of applying DL to a small prospective cohort to detect myofascial pain in the upper trapezius muscle using full B-mode ultrasound videos. This study highlights a data-efficient strategy for testing novel clinical ideas, bridging the gap between early feasibility assessment and large-scale clinical validation.

## 2. Methods and Materials

### 2.1 Participants Cohort and Ultrasound Acquisition

This prospective study was approved by the University of Toronto Institutional Review Board (IRB #: STUDY 22-5584.3). The research project was conducted at the University Center of the Toronto Rehabilitation Institute. Participants were recruited through poster advertisements and provided written informed consent prior to enrollment. Demographic information, including age, sex, and weight, was recorded. Participants were classified into three groups based on the presence of MTrPs: active, latent, or none. The location of MTrPs was determined by manual palpation and marked on the skin with a non-permanent pen by an experienced clinician. Baseline pain level was assessed through visual analog scale, pressure pain threshold (PPT), and brush allodynia testing. A baseline ultrasound scan of the upper trapezius was performed using a commercial ultrasound system (SonixTiuch Q+, Ultrasonix Medical Corporation, British Columbia, Canada), with the transducer aligned parallel to muscle fibers. The scan began at the C7 spinal process and moved linearly towards the acromioclavicular joint at approximately 1cm/s. The transducer captured ultrasound images at 40 frames per second (FPS).

Surface electromyography (sEMG) data were collected using a commercial system (Saga 64, TMSi, Oldenzaal, Netherlands). Electrodes were placed over the marked MTrP location to record muscle activity. Participants performed a shrugging motion with handheld dumbbells of varying weights to activate the muscle, then rested for 2 minutes. They then completed a stretching intervention consisting of three sets of 30-second upper trapezius stretches, with 20-second rest intervals between sets. Following the intervention, participants repeated the shrugging motion with a dumbbell again. The EMG electrode array was then removed, and a second ultrasound scan and pain level measurement were performed afterward. Participants were encouraged to rest as needed during the procedure.

Table 1 illustrates the summary of the study cohort. A total of 24 participants were enrolled, of whom 13 were clinically diagnosed with MTrPs by author DK. For each participant, one pre-intervention and one post-intervention ultrasound video were collected, yielding 48 ultrasound videos for this study. Each video contains an average of 390 frames captured at 40 FPS (10-second duration).

**Table 1.** Study cohort summary.

| Characteristics | Healthy (N=11) | Latent (N=11) | Active (N=2) |
| --- | --- | --- | --- |
| Age | 28.5 (12.5) | 33.7 (11.3) | 25 (1) |
| Gender (N) |  |  |  |
| Male | 8 | 5 | 2 |
| Female | 3 | 6 | 0 |
| *PPT-pre | 50 (15) | 41 (9) | 34 (13) |
| *PPT-post | 52 (18) | 45 (12) | 30 (12) |
| PPT-pre and PPT-post indicate the PPT (Pressure Pain Threshold) value recorded before and after participants performing the stretching intervention. Age, PPT-pre and PPT-post are reported as mean (standard deviation). |  |  |  |

**Table 2.** Parameter settings for Video Masked Autoencoder.

| Patch Size | Encoder Embedded Dimension | Encoder Depth | Encoder Heads | Mask Ratio |
| --- | --- | --- | --- | --- |
| 16 | 384 | 12 | 6 | 0.7 |
| Decoder Embedded Dimension | Decoder Depth | Decoder Heads | MLP Ratio | Tubelet Size |
| 192 | 4 | 4 | 4 | 2 |

### 2.2 Scheme overview

Figure 1 illustrates the developed pipeline for classifying MTrPs using the entire B-mode ultrasound video. A total of 48 ultrasound videos were acquired from 24 participants with either healthy, latent, or active MTrPs of their upper trapezius muscle. Ultrasound videos were preprocessed, and highly overlapping frames were removed from the videos using the Kneedle Algorithm to reduce redundant information and improve performance. Next, the sliding-window approach was applied to resample the preprocessed videos into 404 shorter video clips, each containing 32 frames, which serve as training and testing samples for four-fold cross- validation. Participants were split at the subject level and manually stratified for cross-validation to ensure the models are evaluated on unseen subjects and both positive (latent and active) and negative (healthy) samples are presented in each fold. In each cross-validation iteration, a 2D diffusion model was pretrained on the video frames of the training set with self-supervised learning and incorporated into our proposed diffusion- based classification model, termed Video Diffusion Encoder (VDE), for feature extraction. Both the VDE and baseline models (ResNet, VideoMAE, and SimCLR) are finetuned on the training data and then evaluated on the corresponding hold-out test set. Model performance was assessed using ROC-AUC, F1-score, Accuracy, MCC, and Youden Index.

**Figure 1.**
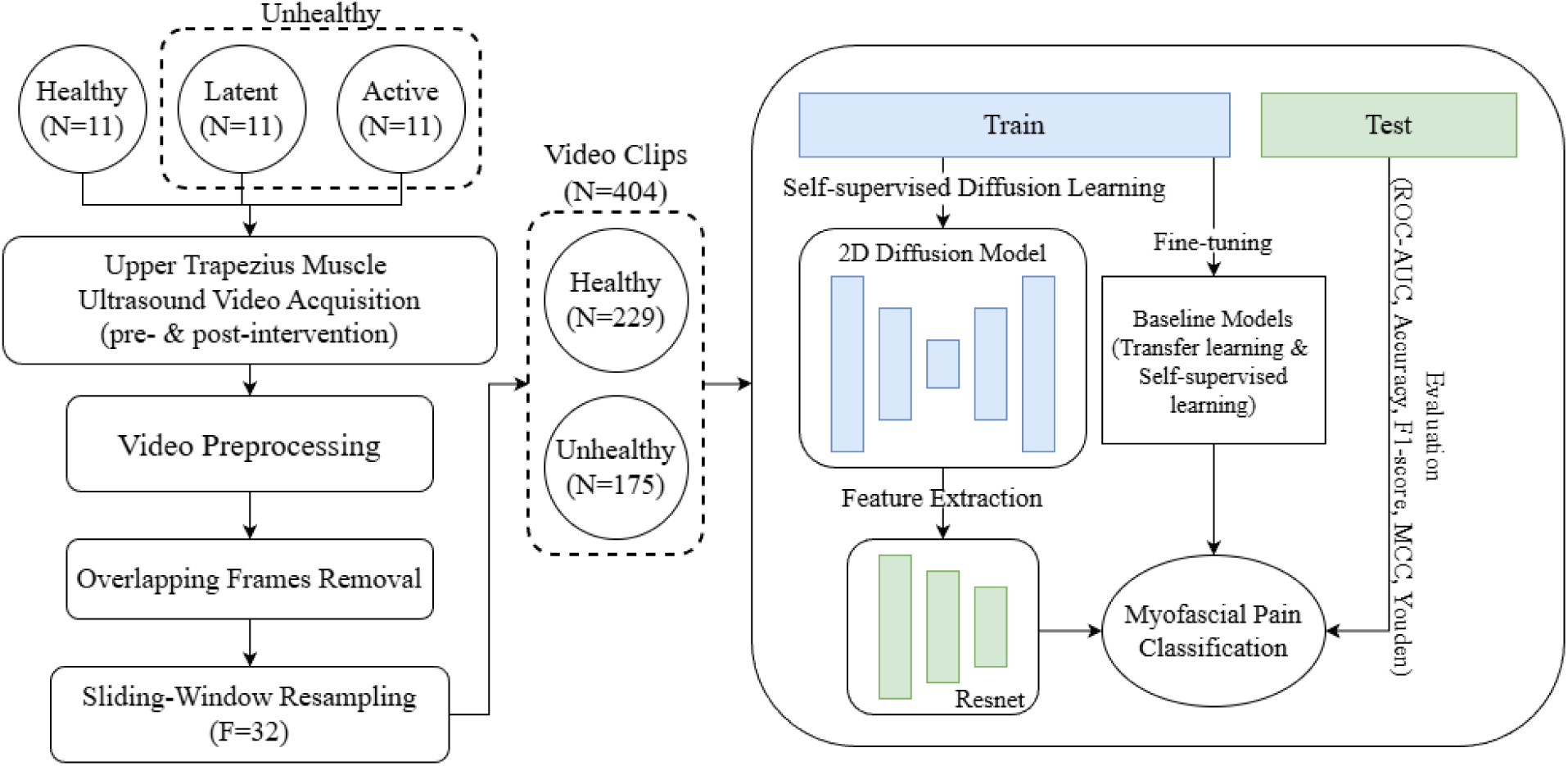
Overview of the pipeline for classifying myofascial pain using B-mode ultrasound video. Ultrasound videos are acquired from participants with either healthy, latent, or active MTrPs. The latent and active participants are grouped into an unhealthy category for binary classification. Ultrasound videos are preprocessed and filtered out high-overlapping frames. The sliding-window approach is employed to resample the preprocessed video into shorter clips for model training. The resampled video dataset is manually stratified and split at the subject level for 4-fold cross-validation. The training samples are used to pretrain the diffusion model with self-supervised learning, and the features produced by the diffusion model are used for MPS classification. The baseline models are also finetuned only on the training set. Performance is evaluated on the hold-out test samples using ROC-AUC, Accuracy, F1-score, MCC, and Youden Index as performance metrics.

### 2.3 Ultrasound Video Preprocessing

Ultrasound videos were processed frame by frame. Each frame was converted to a grayscale image and resized to 224 x 224 pixels for model training. Due to the high frame rate (40 FPS), many consecutive frames were nearly identical, capturing overlapping views of the same muscle region. This redundancy can lead to ineffective learning due to repetitive information. To address this, a previous method proposed by Behr et al. (Behr et al., 2019) is improved and utilized to dynamically determine the threshold for frame removal based on similarity changes across video segments (see Section 2.4 for details). Frame similarity was measured using the complex wavelet structural similarity (CW-SSIM) (Sampat et al., 2009), which ranges from 0 (completely dissimilar) to 1 (identical). Frames with high similarity were removed using a filtering algorithm illustrated in Figure 2.

**Figure 2.**
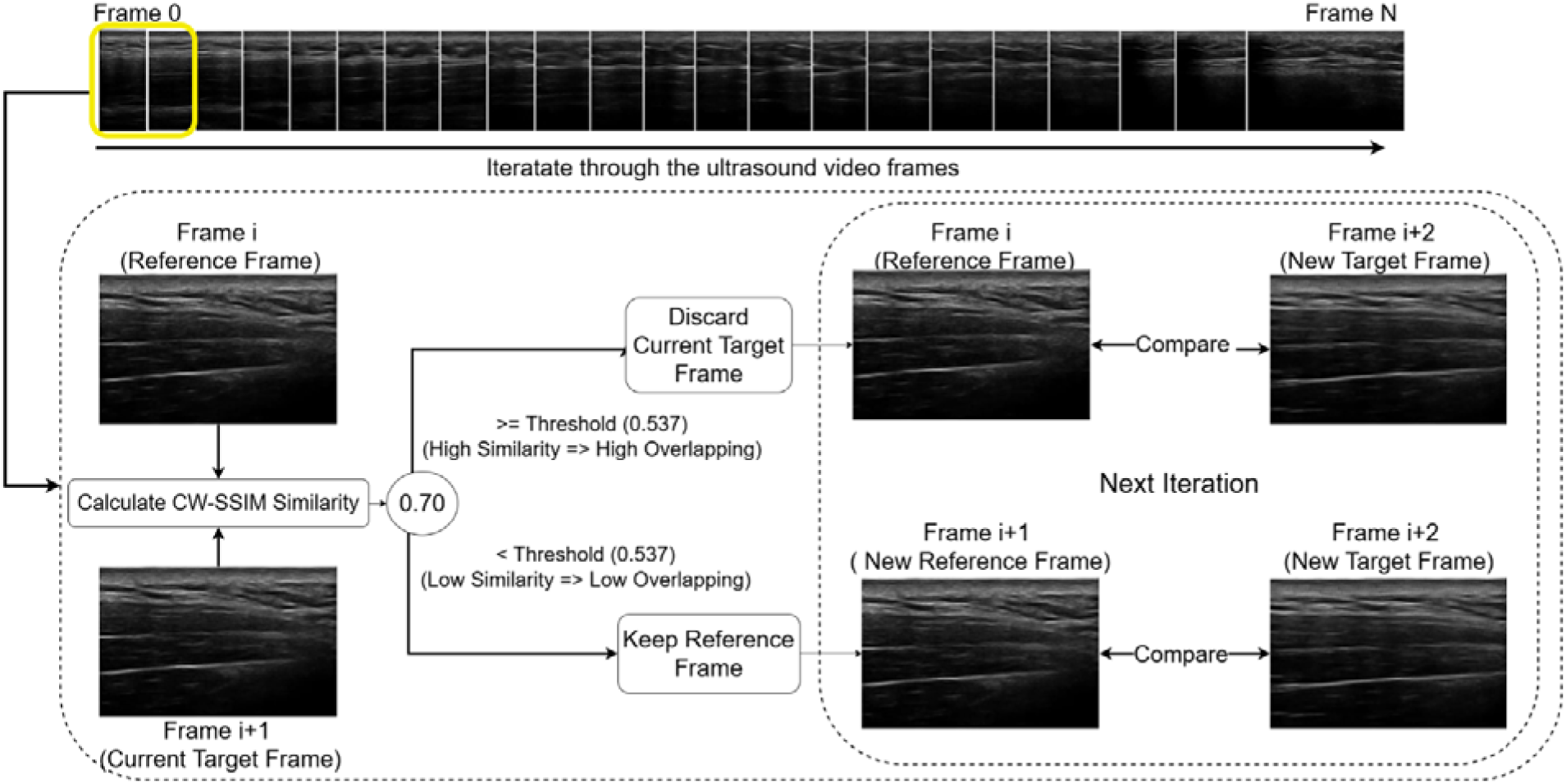
Workflow for removing highly overlapping frames from ultrasound videos using the CW-SSIM similarity value and a threshold. The first frame is set as the reference, and the next as the target. CW-SSIM measures the similarity between them. If the similarity exceeds the threshold (indicating high overlap), the target frame is discarded; otherwise, the reference frame is kept, and the target frame becomes the new reference for the next iteration. The optimal threshold is determined by the improved Kneedle Algorithm (Section 2.4).

The algorithm starts by using the first frame as the reference and the second frame as the target. For each target frame, the CW-SSIM value is calculated relative to the reference frame. If the similarity is below the predefined threshold, the reference frame is kept, and the target frame becomes the new reference. If the similarity exceeds the threshold, indicating high similarity, the target frame is discarded and the reference frame remains unchanged. The optimal threshold is determined by the Kneedle Algorithm (Section 2.4).

### 2.4 Optimal CW-SSIM Threshold Selection Using the Kneedle Algorithm

To determine the optimal threshold for frame selection, the Kneedle Algorithm (Satopaa et al., 2011) is utilized to automatically identify the point where further discarding intermediate frames no longer yields a significant reduction in similarity. This algorithm operates on a curve constructed by computing CW-SSIM values between the first frame of a video and all subsequent frames, capturing how overlapping content changes over time. Initially, the similarity drops as more distinct frames are compared, but eventually, it plateaus. This inflection point, namely the knee, serves as the threshold for deciding which frames to discard. To calculate the knee point, a curve is defined as a finite set of 2D points, where both x and y values are non-negative, then the curve is normalized to fit within a unit square. Next, a difference curve is computed, where the y-values represent the difference between the normalized y- and x-values. This captures the deviation of the curve from the diagonal (i.e., linear growth). To identify where the curve starts to flatten, local maxima are identified in the difference curve, which becomes a candidate for the knee point. A sensitivity parameter S is used to select the knee point from multiple candidates. A smaller value results in earlier selection, while a larger value identifies more candidates before deciding the knee point. Parameter S is set to 1 in this study.

### 2.5. Threshold Curve Calculation

To generate the curve for the Kneedle algorithm, the method proposed by Behr et al. (Behr et al., 2019) is used, which calculates CW-SSIM similarity between the first frame and subsequent frames. However, that approach only considers the start of the video and ignores overlapping variations in later sections. To address this, the previous method is improved by applying a sliding-window approach to the curve calculation to obtain a more robust threshold estimate. Figure 3 shows the curve calculation of the previous (baseline) method and the sliding-window method. In the sliding-window approach, the first frame (i.e., F=1) of each window is treated as the reference, and its similarity is calculated with the next 50 frames. The window then shifts every 20 frames (i.e., F=1, 21, 41, …), and this process continues until the last valid reference (i.e., F=N-50) is reached. The resulting similarity scores from all windows across participants are averaged to form the final similarity curve. The knee point of this curve is then calculated through the Kneedle algorithm and determined as the optimal threshold for frame selection. Unlike the previous method, the improved approach captures overlapping differences throughout the entire video, not just at the beginning, leading to a more accurate and representative threshold for preprocessing.

**Figure 3.**
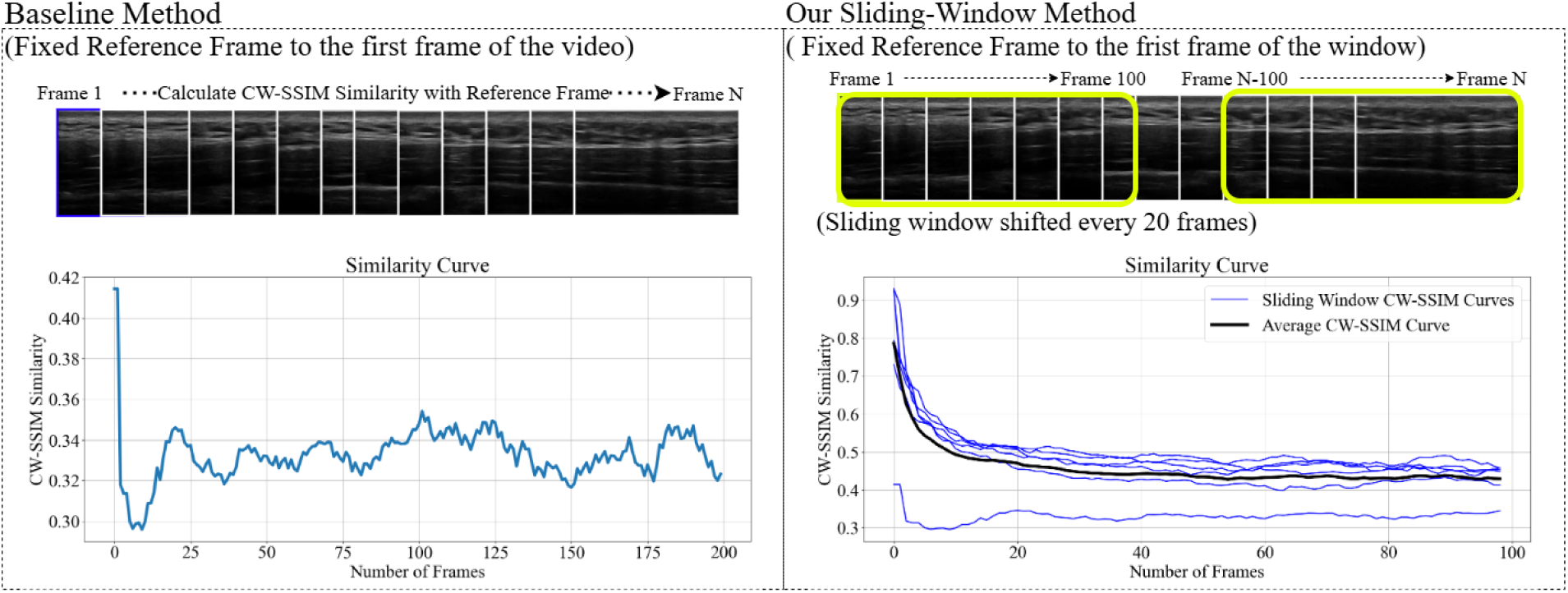
Illustration of similarity curve calculation. A) In the prior method proposed by Behr et a,l(Behr et al., 2019) the first frame of the video is used as a fixed reference, and the similarity is computed between it and all subsequent frames to generate the similarity curve. B) In our sliding-window approach, the reference frame shifts every 20 frames, and similarity is calculated between each reference frame and its next 100 frames. The final similarity curve is obtained by averaging the similarity curves from all windows.

### 2.6 Sliding Window Resampling

To increase the number of training samples, a sliding-window resampling strategy is applied to the preprocessed ultrasound videos. Each video is segmented into clips of 32 consecutive frames using a step size of 1. For videos containing fewer than 32 frames after preprocessing, frames were reversed and appended to the original sequence to create a looping clip of 32 frames. Because the looping clip preserves all original frames, only a single clip was generated for these videos.

### 2.7. Baseline Models

Three representative video classification approaches were selected as the baseline for MTrPs classification, including 3D ResNet models of different depths (i.e., ResNet-50, ResNet-101), a lightweight Video Masked Autoencoder (VideoMAE), and SimCLR contrastive learning with a 3D ResNet-50 backbone (Chen et al., 2020; Tong et al., 2022).

To address the limited sample size and improve generalizability, transfer learning is employed using the large-scale Kinetics-400 dataset (Kay et al., 2017), which contains 400 human action classes with 400 videos per class. The ResNet models were initialized with weights pretrained on Kinetic-400. During finetuning, only the final ResNet block and the classification head are updated using the ultrasound video dataset, while earlier layers were frozen to mitigate overfitting.

For self-supervised baselines, VideoMAE, is pretrained on the ultrasound video clips by reconstructing the masked image patches, allowing the model to learn ultrasound-specific representations that generalize well to small datasets. After pretraining, the decoder is discarded, the encoder is frozen, and linear probing is applied by finetuning only the final classification layer for MPS classification. SimCLR contrastive learning is conducted using a 3D ResNet-50 backbone to learn video representations from ultrasound clips. Following contrastive pretraining, the encoder is frozen, and linear probing is applied for MPS classification.

### 2.8 Video Diffusion Encoder

To address the limited sample size and improve generalizability, a novel approach called Video Diffusion Encoder (VDE) is developed that leverages self-supervised learning via a diffusion model to extract robust and meaningful features from ultrasound video frames for video classification. An overview of the VDE pipeline is shown in Figure 4.

**Figure 4.**
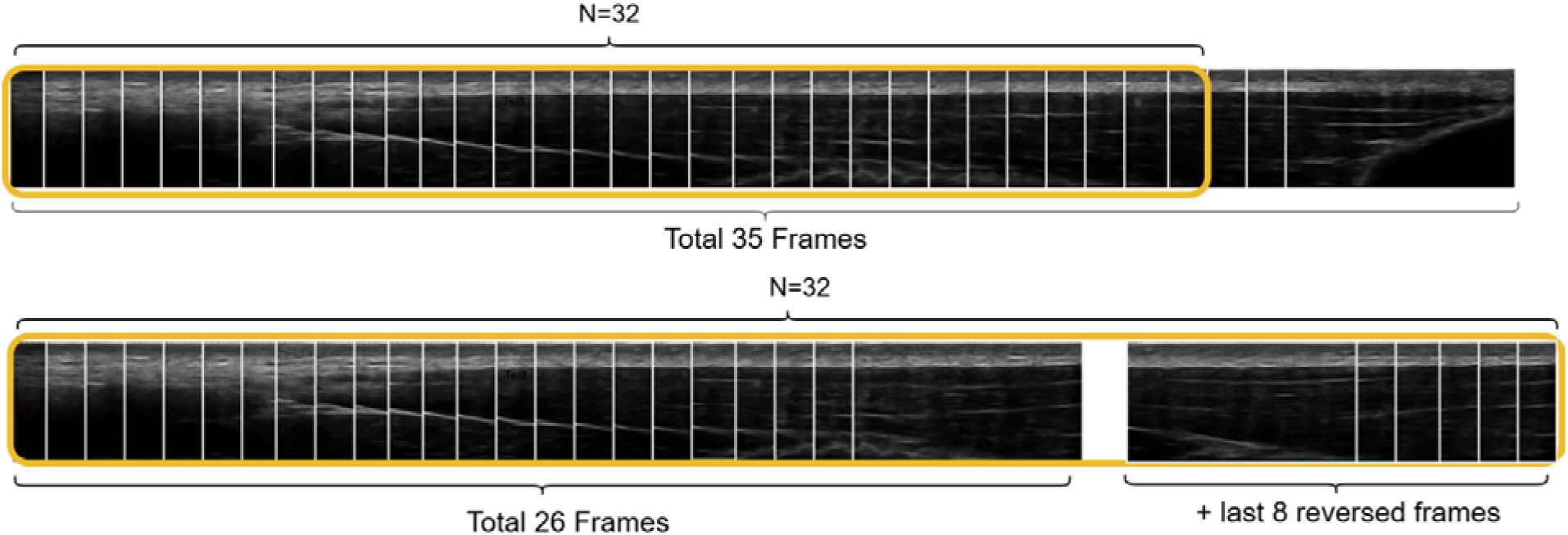
Illustration of the sliding-window resampling strategy. Videos with more than 32 frames are segmented using a 32-frame sliding window with a step size of 1. Videos with fewer than 32 frames are extended by appending reversed frames to form a single 32-frame looping clip for training and validation.

**Figure 5.**
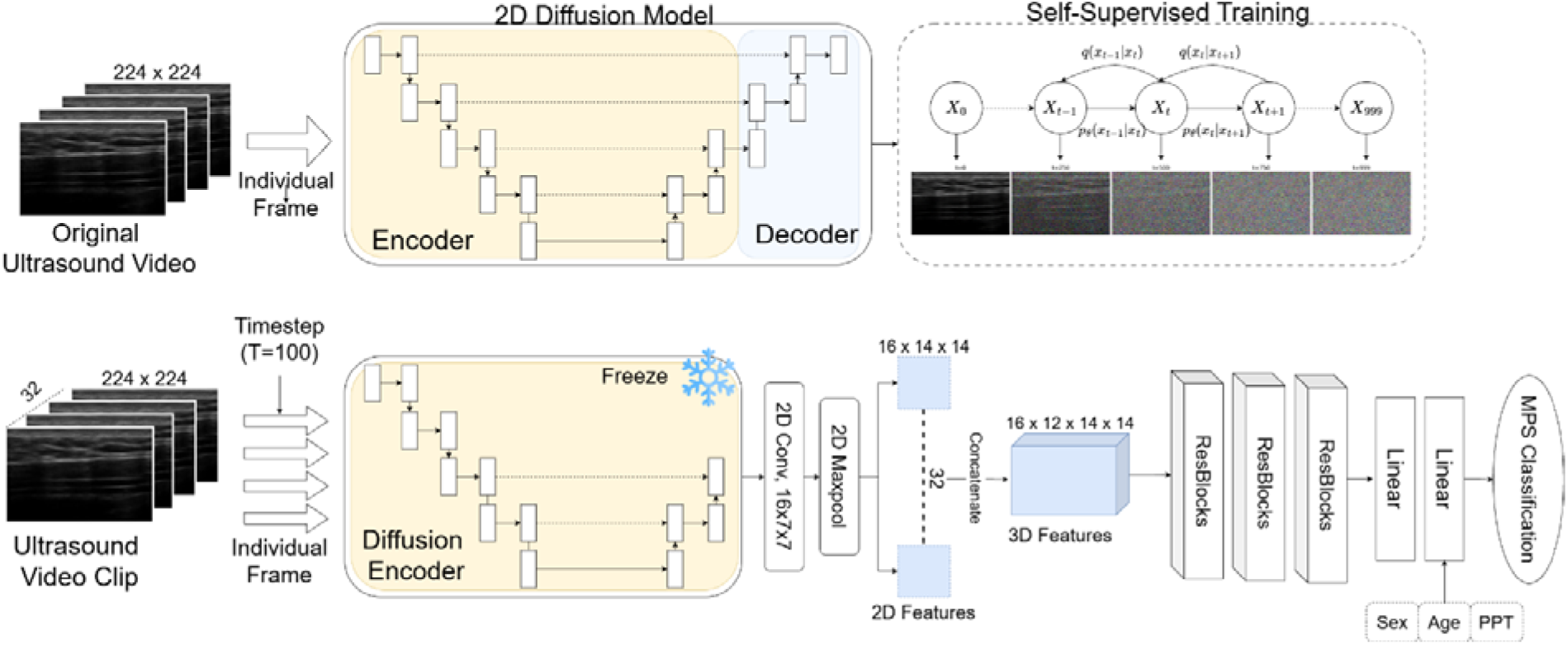
Overview of the Video Diffusion Encoder (VDE). First, a 2D diffusion model is pretrained on original ultrasound video frames using self-supervised diffusion learning with various augmentations. After pretraining, the model parameters are frozen. Each frame in the video clip is passed through the encoder part of the diffusion model (up to the middle of the up-sampling block) at a fixed timestep to extract 2D features. These features are concatenated along the temporal axis to form a 3D representation, which is then processed by several 3D ResNet blocks, followed by linear layers for MPS prediction. Additionally, demographic (e.g., age, gender) and clinical (e.g., PPT) information is concatenated with flattened video features before the second linear layer.

First, a 2D diffusion model is pre-trained on frames of the original (unfiltered) ultrasound videos in the training folds (N=3). Once trained, its weights are frozen, and the encoder portion is used to extract frame- wise features from video clips resampled via the sliding-window strategy. Each frame is passed through the encoder part with a fixed timestep, producing frame-level 2D features. These frame-level features are processed with a convolution layer and a max pooling layer to reduce spatial dimensions, then concatenated across time to produce a 3D feature volume. The 3D representation is fed into multiple 3D ResNet blocks for further feature extraction, followed by linear layers for binary classification. Additionally, demographic and clinical information, including sex, age, and PPT value, was incorporated into the model by converting it to embedding vectors and concatenating with video features before the second linear layer. The diffusion encoder is defined as the first 13 blocks of the U-Net backbone in the diffusion model, following best practices from prior work (Mukhopadhyay et al., 2024; Xiang et al., 2023). The full backbone of the diffusion model consists of 18 blocks: 8 down-sampling, 2 middle, and 8 up-sampling blocks. Based on prior findings that optimal features can be extracted from the middle of the up-sampling path along with a low timestep, blocks 1–13 is used for feature extraction, and while the diffusion model is trained with a total of 1000 timesteps, the inference for acquiring 2D frame-wise features is performed using a timestep of 100, as determined by our ablation study. Each residual block contains 32, 64, and 128 channels, respectively, and the output dimension of the first linear layer is set to 128. The PPT feature represents the average PPT value of both pre- and post-intervention, and the size of the embedding vector for each demographic and clinical feature is set to 4.

### 2.9 Training Procedure

We performed stratified four-fold cross-validation at the subject level. All videos from the same participant, including pre- and post-intervention scans, were assigned to the same fold to prevent data leakage. The diffusion model was pretrained exclusively on ultrasound video frames from the three training folds, ensuring no overlap with the evaluation fold. Diffusion pretraining uses an initial learning rate of 0.0001, a cosine beta scheduler, the Adam optimizer, and Mean Squared Error (MSE) loss. To enhance data diversity and mitigate overfitting, data augmentations, such as random flipping, random rotation, random translation, and random Gaussian blurring, were applied to ultrasound images. The diffusion encoder was frozen during supervised classification training,

For baseline ResNet models, only the final ResNet block and classification head were finetuned on ultrasound videos, with a learning rate of 0.00001, AdamW optimizer, and Binary Cross-Entropy (BCE) loss. The classification head contains three linear layers with a dropout rate of 0.5 applied between layers to mitigate overfitting.

The VideoMAE baseline was pretrained using self-supervised masked reconstruction with a learning rate of 0.0001, the AdamW optimizer, and Mean Squared Error (MSE) loss. For classification, we discarded the decoder and adopted a linear probing method, finetuning only the final linear layer after the frozen encoder for MPS classification. The SimCLR contrastive learning was applied with a learning rate of 0.001, AdamW optimizer, and NT-Xent loss with a temperature of 0.5. Two different augmentation pipelines were used for contrastive learning. The first pipeline contains random rotation, random Gaussian blurring, and random shearing. The second pipeline contains random translation, random Gaussian blurring, and random rotation. After contrastive pretraining, the linear probing, where the backbone ResNet-50 was frozen and only the classification head was finetuned, was applied for MPS classification.

All baseline models were finetuned for 20 epochs with the same learning rate (0.0001), the AdamW optimizer, and Binary Cross Entropy (BCE) Loss, for MPS classification. Augmentations, including random flipping, rotation, translation, and Gaussian blurring, were applied. All trainings were performed on an NVIDIA GeForce RTX 3090 GPU.

### 2.10 Evaluation

Models were evaluated on the validation set to assess their performance on unseen subjects’ ultrasound videos. The best-performing models on the validation set were saved and used for final evaluation. Each model underwent 5 runs of four-fold cross-validation to compute average performance metrics. Evaluation metrics include ROC-AUC, F1-score, Accuracy, MCC, and Youden Index. As the model outputs probability from 0 to 1 for binary classification, the decision boundary is determined by the threshold that maximizes the Youden Index. MCC, which accounts for true positive (TP), true negative (TN), false positive (FP), and false negative (FN), is considered a more balanced and reliable metric for binary evaluation (Eq. 6). It produces a value range from −1 to +1, where +1 indicates perfect classification and −1 indicates complete disagreement.

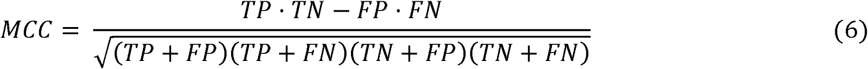

The Youden Index (Schisterman et al., 2008) was calculated to evaluate binary classification performance and determine the optimal decision boundary for binary classification. By balancing sensitivity and specificity, the Youden Index is widely used in medical diagnostics to identify the threshold that best separates positive and negative classes. Its value ranges from 0 (random prediction) to 1 (perfect prediction).

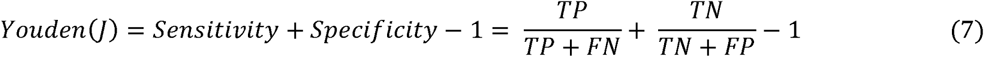

Predicted probabilities for individual video clips are further aggregated by averaging the predictions from the same video and the same subject to generate video-level and subject-level probabilities. Model performance is evaluated and reported at the clip, video, and subject levels using the same set of evaluation metrics. The Wilcoxon signed-rank t-test was used to compare the MCC score between VDE and baseline models at different aggregation levels.

To address the small number of active MTrPs subjects (N=2) relative to latent MTrPs subjects (N=11), a latent-only analysis was conducted to compare the performance between latent-only groups versus latent and active groups using ROC-AUC at clip, video, and subject levels. This analysis provides a more robust assessment of model prediction for latent subjects that are not affected by active subjects. The analysis was conducted by removing active subjects during training and evaluation for latent-only groups, and the performance between the two groups (latent-only and latent with active) was evaluated and compared only on latent subjects, even though the latent and active groups were trained on active subjects. The Wilcoxon signed-rank t-test was used to compare the ROC-AUC score of two groups.

### 2.11 Model Interpretability

To interpret the prediction generated by the VDE model, Gradient-weighted Class Activation Mapping (Grad-CAM) (Selvaraju et al., 2017) was employed to visualize image regions that most strongly influenced model outputs. Gradients from the final 3D ResNet block were used to compute importance weights for the corresponding feature maps. These weighted feature maps were then up-sampled to the input image resolution and overlaid on the original images to generate interpretable heatmaps highlighting salient regions driving the model’s predictions.

## 3 Results

### 3.1 Preprocessed and Sliding-Window Resampling Result

Using 50 frames with a step size of 20 for threshold curve calculation, the average number of frames per ultrasound video dropped from 3900±059 to 370± 10. Sliding-window resampling produced a total of 404 video clips per cross-validation iteration, including an average of 303 training and 101 validation clips. Among the 404 video clips, 229 were labeled healthy and 175 unhealthy.

### 3.2. Prediction performance

Table 3 summarizes the average performance of the baseline models and proposed video diffusion encoder (VDE) model using F1-score, Accuracy, MCC, and Youden Index at the clip-, video-, and subject-levels. Figure 6 presents the corresponding ROC curves.

**Figure 6:**
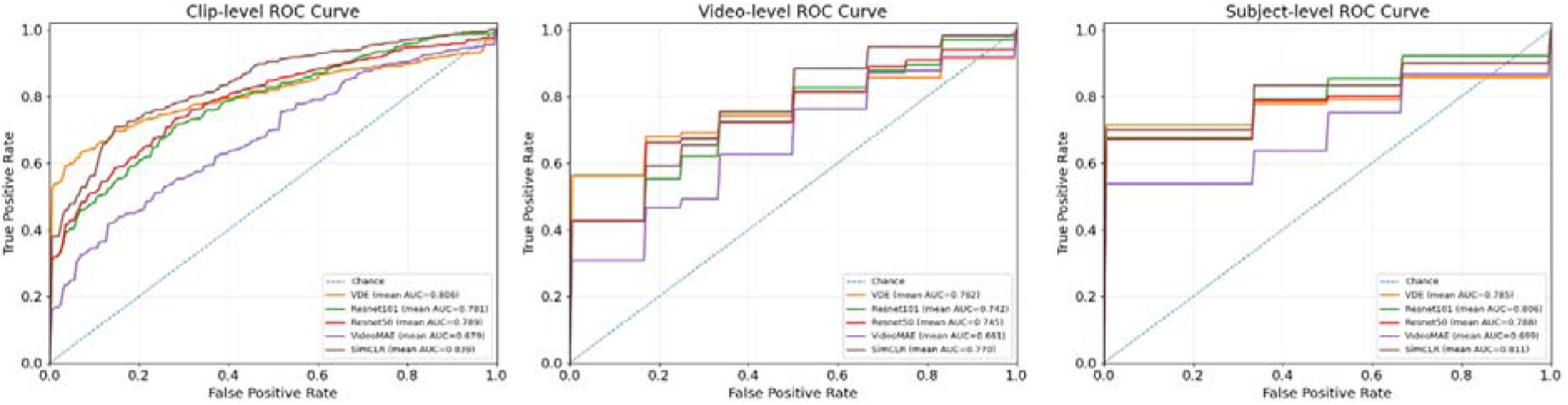
Average ROC Curve of all models with different aggregation levels (left to right: clip-level, video-level, subject-level).

**Table 3:** The average clip-level, video-level, and subject-level performance of each model. Reported with mean and 95% confidence intervals.

| Models | AUC | F1-score | Accuracy | MCC | Youden | p-value |
| --- | --- | --- | --- | --- | --- | --- |
| **Clip Level (N=404)** |  |  |  |  |  |  |
| Resnet-50 | 0.79 (0.73-0.83) | 0.72 (0.65-0.80) | 0.77 (0.72-0.82) | 0.56 (0.48-0.64) | 0.56 (0.48-0.63) | 0.08 |
| Resnet-101 | 0.78 (0.73-0.83) | 0.71 (0.63-0.77) | 0.75 (0.69-0.81) | 0.53 (0.45-0.62) | 0.53 (0.45-0.61) | **0.02** |
| VideoMAE | 0.76 (0.70-0.83) | 0.71 (0.60-0.82) | 0.76 (0.68-0.84) | 0.57 (0.44-0.70) | 0.57 (0.43-0.70) | **0.02** |
| SimCLR | **0.84 (0.77-0.89)** | **0.76 (0.68-0.83)** | 0.83 (0.80-0.86) | 0.60 (0.51-0.69) | 0.61 (0.51-0.69) | 0.31 |
| VDE (T=100) | 0.81 (0.71-0.89) | 0.75 (0.62-0.85) | **0.85 (0.78-0.89)** | **0.66 (0.54-0.77)** | **0.64 (0.52-0.75)** | - |

| Video Level (N=48) |  |  |  |  |  |  |
| --- | --- | --- | --- | --- | --- | --- |
| Resnet-50 | 0.74 (0.69-0.79) | 0.75 (0.69-0.80) | 0.77 (0.74-0.80) | 0.60 (0.54-0.65) | 0.57 (0.50-0.65) | 0.36 |
| Resnet-101 | 0.74 (0.70-0.78) | 0.76 (0.72-0.81) | 0.78 (0.74-0.81) | 0.61 (0.55-0.67) | 0.57 (0.51-0.63) | 0.62 |
| VideoMAE | 0.66 (0.58-0.73) | 0.69 (0.60-0.77) | 0.71 (0.66-0.77) | 0.51 (0.42-0.60) | 0.47 (0.37-0.56) | 0.31 |
| SimCLR | 0.77 (0.70-0.83) | 0.76 (0.70-0.82) | 0.77 (0.72-0.82) | 0.57 (0.47-0.66) | 0.55 (0.45-0.65) | 0.15 |
| VDE (T=100) | **0.77 (0.66-0.86)** | **0.76 (0.65-0.86)** | **0.81 (0.75-0.87)** | **0.66 (0.55-0.77)** | **0.65 (0.52-0.76)** | - |
| Subject Level (N=24) |  |  |  |  |  |  |
| Resnet-50 | 0.74 (0.72-0.85) | 0.85 (0.81-0.89) | 0.84 (0.80-0.88) | 0.73 (0.65-0.80) | 0.70 (0.63-0.79) | 0.60 |
| Resnet-101 | 0.80 (0.75-0.87) | 0.83 (0.63-0.77) | 0.83 (0.69-0.81) | 0.72 (0.45-0.62) | 0.69 (0.45-0.61) | 0.72 |
| VideoMAE | 0.69 (0.58-0.73) | 0.69 (0.60-0.82) | 0.76 (0.68-0.84) | 0.60 (0.44-0.70) | 0.57 (0.43-0.70) | 0.73 |
| SimCLR | **0.81 (0.72-0.88)** | **0.84 (0.76-0.90)** | 0.86 (0.81-0.91) | **0.77 (0.68-0.85)** | 0.73 (0.63-0.83) | 0.26 |
| VDE (T=100) | 0.79 (0.66-0.90) | 0.83 (0.71-0.91) | **0.86 (0.79-0.91)** | 0.66 (0.54-0.77) | **0.73 (0.60-0.85)** | - |

At the clip level, the VDE with a diffusion timestep of 100 outperformed transfer learning-based ResNet and VideoMAE with a statistically significant difference (p<0.05) and achieved comparable performance with SimCLR. VDE achieves an AUC of 0.81 (95% CI: 0.71–0.89), accuracy of 0.85 (95% CI: 0.78–0.89), F1- score of 0.75 (95% CI: 0.62–0.85), MCC of 0.66 (95% CI: 0.54–0.77), and Youden Index of 0.64 (95% CI: 0.52–0.75). SimCLR performed the best at AUC and F1-score, with an AUC of 0.84 (95% CI: 0.77–0.89), accuracy of 0.83 (95% CI: 0.80–0.86), F1-score of 0.76 (95% CI: 0.68–0.83), MCC of 0.60 (95% CI: 0.51– 0.69), and Youden Index of 0.61 (95% CI: 0.51–0.69).

At the video level, VDE consistently outperformed the baseline models, yielding an AUC of 0.77 (95% CI: 0.66–0.86), accuracy of 0.81 (95% CI: 0.75–0.87), F1-score of 0.76 (95% CI: 0.65–0.86), MCC of 0.66 (95% CI: 0.55–0.77), and Youden Index of 0.65 (95% CI: 0.52–0.76). At the subject level, VDE and SimCLR showed comparable performance. VDE achieved an AUC of 0.79 (95% CI: 0.66–0.90), accuracy of 0.86 (95% CI: 0.79–0.91), F1-score of 0.83 (95% CI: 0.71–0.91), MCC of 0.66 (95% CI: 0.54–0.77), and Youden Index of 0.73 (95% CI: 0.60–0.85). Neither video-level nor clip-level performance was statistically significant (Table 3).

### 3.3 Latent Trigger Point-only Analysis

Table 4 compares model performance between latent-only groups and combined latent and active groups across each fold and on average. Overall, the AUC scores for the latent-only groups were not significantly different from the combined groups. Notably, in fold 3, the latent-only group shows higher AUC score (clip- level: 0.954, 95% CI: 0.91-0.98; video-level: 0.87, 95% CI: 0.83-0.95, subject-level: 0.95, 95% CI: 0.91-0.98) compared to the combined group (clip-level:0.748, 95% CI: 0.641-0.870; video-level: 0.72, 95% CI :0.60- 0.81; subject-level: 0.74, 95% CI: 0.64-0.87).

**Table 4.**
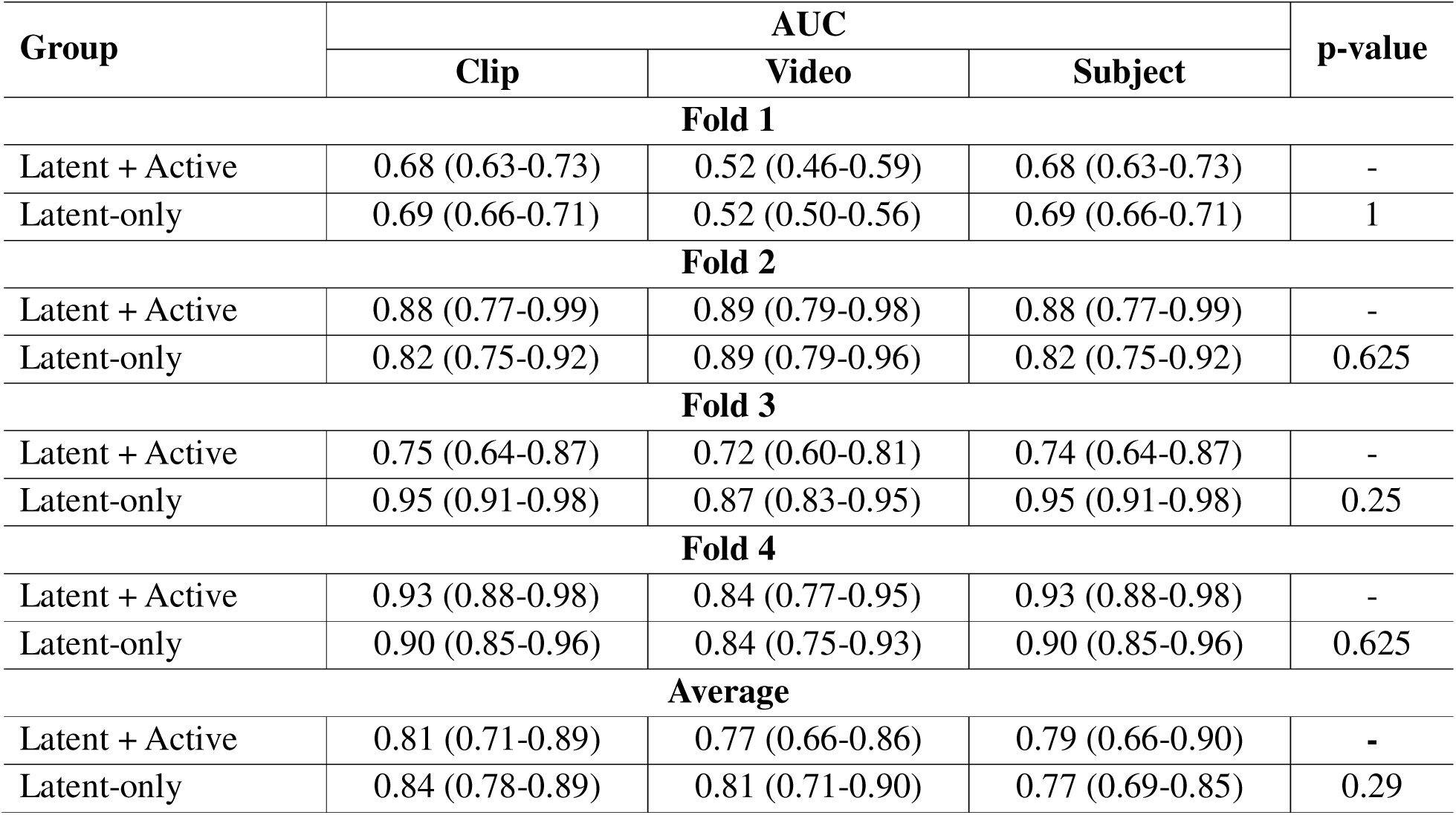
Performance comparison between latent trigger point-only and the combined latent and active groups across each fold and on average. Values reported as mean and 95% confidence intervals (CIs).

### 3.4 Performance Across Different Frame Sizes

Figure 7 shows the effect of frame number on the ResNet-50 model. Using 32 frames yielded the highest AUC of 0.803 (95% CI: 0.75-0.86). Performance decreased slightly across other frame counts: 40 frames produced the second-best AUC of 0.801 (0.74–0.86), while 24 and 48 frames yielded slightly lower results.

**Figure 7.**
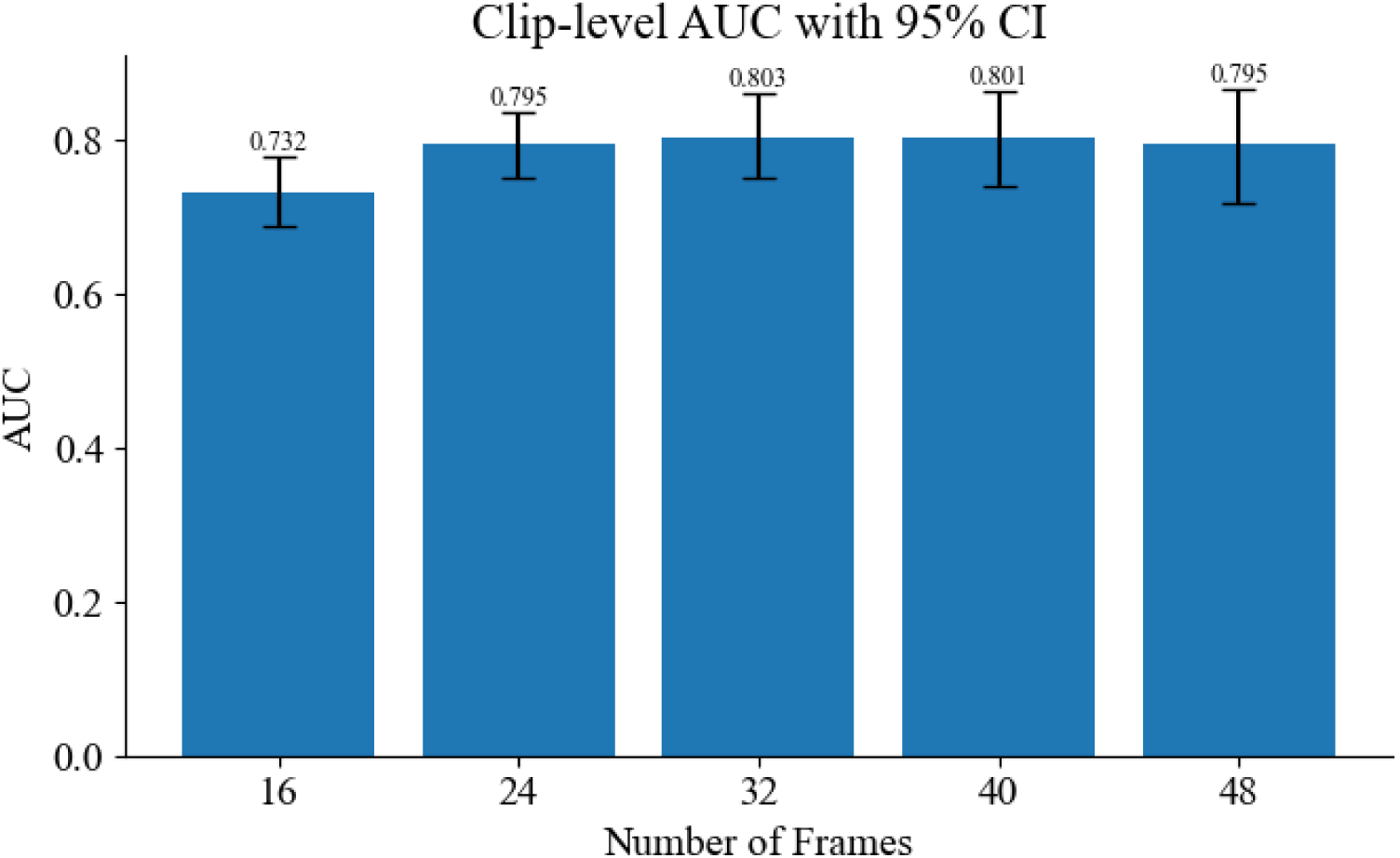
Classification performance across different frame sizes using Resnet-50.

### 3.5 Performance Difference Across Diffusion Timesteps

Figure 8 illustrates the clip-level AUC difference of the VDE model across different timesteps used during classification. Overall, a timestep of 100 yields the best performance, with an AUC of 0.811 (95% CI: 0.710- 0.895), followed by a timestep of 50 with an AUC of 0.796 (95% CI: 0.732-0.862).

**Figure 8:**
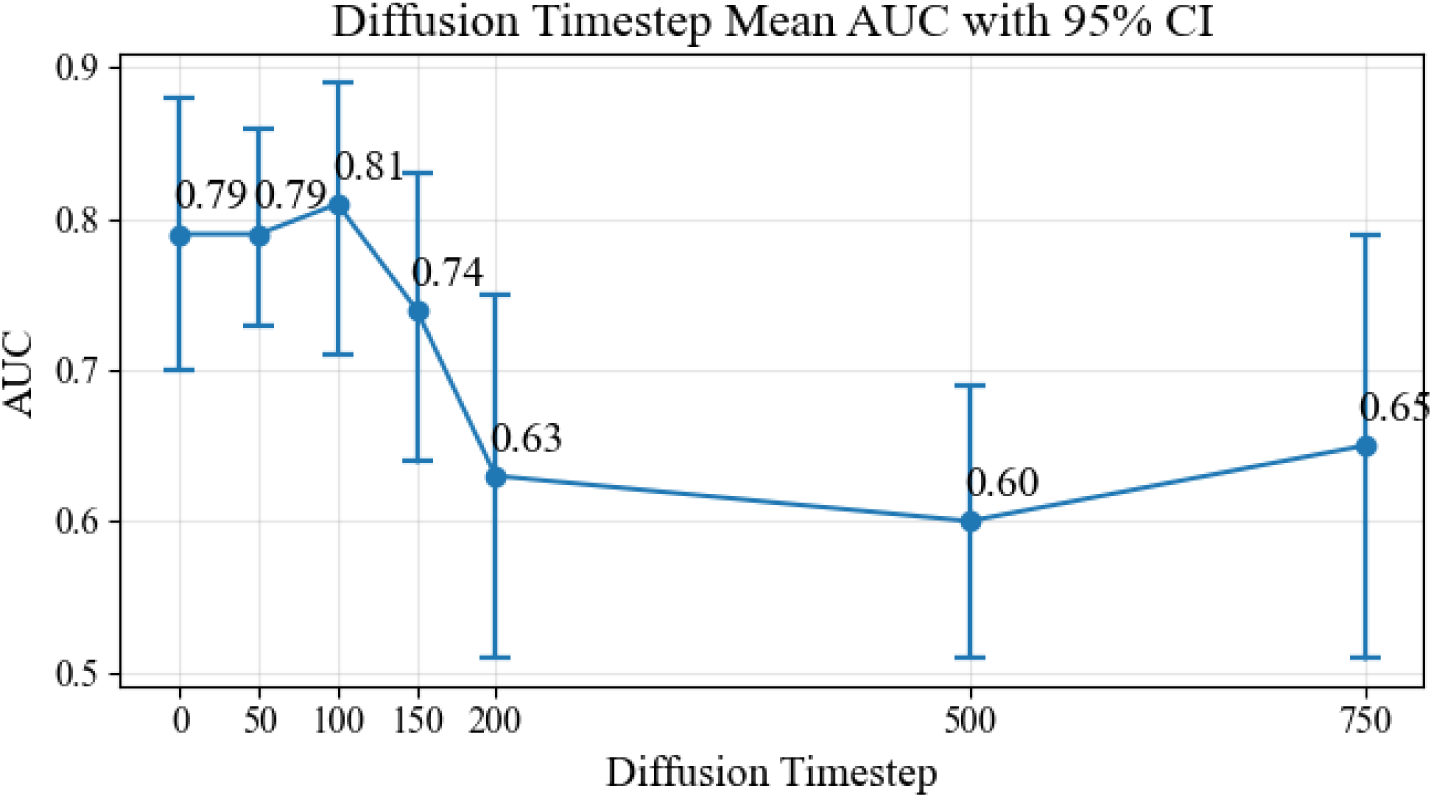
Classification performance differences across diffusion timesteps

### 3.6 Interpretability Analysis

Figure 9 shows the Grad-CAM visualization used to interpret VDE model predictions across classes and pre- and post-intervention conditions. The highlighted regions correspond to image areas that contributed most strongly to the model’s decision. Across all classes and intervention states, the model often attends to superficial soft tissue and fascia regions (bright region), where muscle tissue (dark region) is less frequently emphasized.

**Figure 9.**
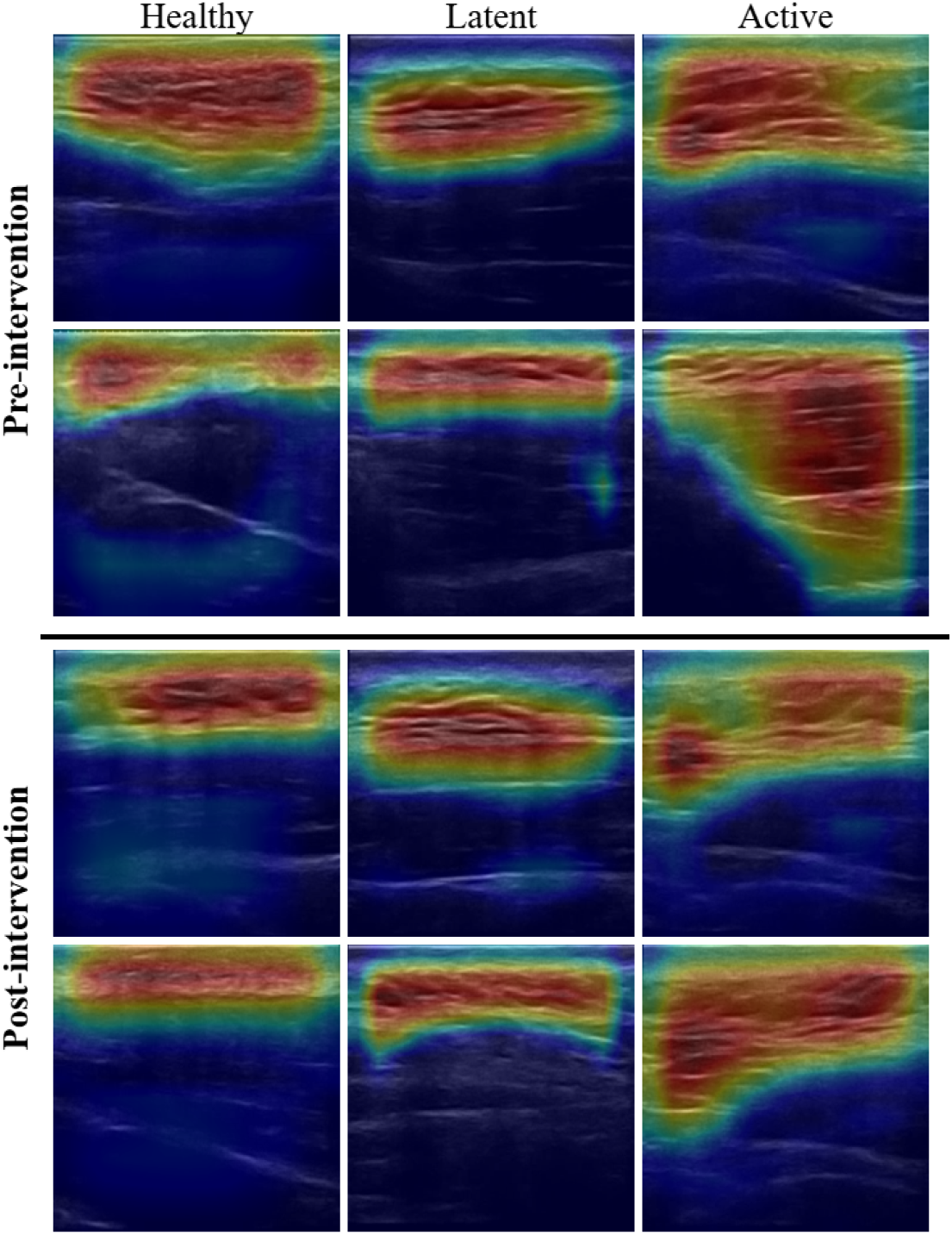
Grad-CAM-based interpretability analysis of the VDE model across classes and pre- and post-intervention conditions.

## 4 Discussion

In this study, we employed deep learning approaches to investigate the feasibility of using full B-mode ultrasound videos for identifying MPS in a prospective cohort. Our approach has several advantages. First, it eliminates subjectivity from manual palpation, as predictions are based solely on objective image data without patient or clinician input. Second, it removes the need for manual frame selection (Behr et al., 2019; Koh et al., 2023; Shomal Zadeh et al., 2023), which are often subjective and varies among observers. Automating this step improves consistency and reduces the risk of missing relevant information, especially since hypoechoic MTrPs do not always present in a single frame. Third, it allows the model to capture subtle motion patterns, tissue dynamics, and temporal changes that are often lost in static frames or cropped clips. Finally, frame-by-frame review is not scalable in clinical practice, whereas a model that processes entire recordings offers a more efficient and practical solution for real-world use. As demonstrated by the experiments, the VDE approach achieved strong and consistent performance across multiple aggregation levels (Table 3.), highlighting the potential of automating MTrPs detection using full B-mode ultrasound video and suggesting that even within the constraints of a small, prospectively collected dataset, deep learning can extract informative features from ultrasound to support MTrP detection. It also demonstrates the potential of developing video-based biomarkers to further differentiate active versus latent MTrPs. Although VDE only showed statistically significant performance improvements at the clip level and comparable performance to contrastive learning, it consistently achieved strong overall performance across all evaluation metrics at different aggregation levels. Furthermore, the relatively small cohort size may limit statistical power, thereby reducing the ability to fully distinguish differences in predictive performance among the models.

During ultrasound video acquisition, the high frame rate often results in highly overlap consecutive frames, introducing redundant information that can increase computational resource usage and degrade model performance. To address this, the previously proposed method (Behr et al., 2019) is improved and utilized to remove highly similar frames. In their method, the similarity curve used for threshold determination is computed by comparing all frames to the first frame. In contrast, our sliding-window approach measures frame similarity across segments of the video, better capturing variations in overlap throughout its duration and enabling a more accurate and adaptive threshold for frame removal, which is essential because setting the threshold too high retains redundancy, while too low discards useful data. The improved method provides better data quality and training efficiency. This approach can also be applied to other imaging modalities where frame redundancy is a concern. The sliding-window parameters for threshold calculation (window size = 50, step size = 20 frames) are selected to achieve an average of approximately 32 remaining frames per video after redundancy removal, ensuring compatibility with GPU memory constraints while retaining sufficient temporal information. This configuration yielded satisfactory performance in our experiments. Further ablation studies can be conducted to investigate the impact of different threshold-calculation settings and sliding-window resampling strategies on model performance. Other parameters, such as the number of frames per clip and the diffusion timestep, were determined through ablation studies, where 32 frames per clip and a diffusion timestep of 100 yielded the best performance and were therefore used for final evaluation.

To maximize the utility of a small prospective study cohort, self-supervised diffusion learning is leveraged for feature representation learning and allows analysis to be performed under the condition of a limited dataset. Based on the result of cross-validation, this approach outperforms transfer learning ResNet and self-supervised videoMAE across all metrics and achieves performance comparable to SimCLR (Table 3), demonstrating that the multi-level denoising process can effectively capture meaningful features from ultrasound images and improve video classification, highlighting the potential of applying diffusion learning for ultrasound analysis. Additionally, the denoising process may help mitigate common ultrasound artifacts such as backscatter and noise by isolating noise from clinically relevant features.

Despite the large domain gap between kinetic-400 and our ultrasound dataset, the transfer learning-based model still achieves reasonable performance after finetuning. This aligns with prior findings that pre-training on natural images can still provide useful low- and mid-level feature representations when adapted to medical images (Zu et al., 2025). We intentionally avoided large-scale domain-specific pretraining models (e.g., ultrasound foundation models) to highlight the benefit of self-supervised diffusion learning for a small ultrasound video dataset, as domain-specific pretraining would obscure the impact of our proposed methodology.

The Grad-CAM method was used to interpret the VDE model prediction from ultrasound videos. The feature map visualization (Figure 8) shows that the model consistently focused on superficial soft tissue and fascia (bright) region while largely ignoring muscle tissue (dark). This suggests that, on ultrasound, myofascial pain may be identified in the superficial soft tissue and fascial region rather than within the muscles. Although trigger points typically reside in muscle tissue, this is physically plausible, as fascial dysfunction can contribute to MPS through multiple potential mechanisms, including densification, fascial thickening, neuroinflammation, and chronic fascial changes (Gromakovskis 2025). However, saliency methods like Grad-CAM can be spatially coarse and influenced by high-contrast boundaries; therefore, it should be interpreted cautiously. Nonetheless, these findings support the conclusion that the model relied on clinically meaningful image cues rather than arbitrary noise, with salient regions closely aligning with clinician-identified diagnostic areas.

Since active and latent MTrPs may exhibit different symptoms and imaging appearances, they should ideally be analyzed separately. Due to limited sample sizes, both types were grouped into a single MPS category for classification. To assess potential bias from active subjects, a latent-only analysis was conducted. Results show no significant performance differences between the latent-only and combined groups, indicating that the model primarily relies on general image features common to both active and latent MTrPs, and, therefore, removing active subjects has minimal impact.

Several limitations should be acknowledged. First, the small prospective cohort introduces inherent challenge for early-stage studies. To mitigate the risk of overfitting and obtain a reliable estimate of model performance, we adopted subject-level cross-validation and reported performance at the clip, video, and subject levels. In addition, we leveraged self-supervised learning and transfer learning from large-scale video datasets to obtain generalizable video features prior to ultrasound-specific fine-tuning. This supports the feasibility and innovation of our data-efficient approach as a proof-of-concept for evaluating novel ideas before scaling larger studies. Second, due to the limited number of MPS participants, active and dominant latent MPS cases were grouped into a single unhealthy category. But because active and latent MPS differ in pain-related physiology and the dataset is dominated by latent cases, this grouping may overgeneralize the ability to identify active myofascial pain. However, the small number of active cases precluded a separate analysis; therefore, a latent-only analysis was performed to assess predictive ability in latent subjects, and conclusions are limited to the model’s ability to distinguish MPS patients from healthy participants. Third, the current labels are determined by manual palpation. Although palpation was performed by an experienced clinician, this approach may still introduce subjective bias due to the inherent variation of manual assessment. Since the model was trained based on these labels, our reported performance may represent an upper bond of true performance. Finally, this study lacks an external validation cohort, which limits the generalizability of our findings across different clinical settings and populations. Such cohorts are particularly difficult to obtain in the context of prospective research, where comparable datasets are scarce and multi-site recruitment requires significant time and coordination. As a result, opportunities for cross-site validation or benchmarking against existing work are currently limited. Nevertheless, the primary goal of this study was not large-scale generalization, but rather to demonstrate that deep learning can be feasibly applied to a small, prospectively collected cohort as an innovative proof-of-concept for identifying myofascial pain. The promising results presented here provide a foundation for future multi-center collaborations and large-scale validation.

## 5 Conclusion

This study demonstrates the feasibility of applying deep learning to identify myofascial pain in the upper trapezius muscle using full B-mode ultrasound videos within a prospective cohort. Recognizing that prospective studies face natural challenges of small sample sizes and slow enrollment, we introduced a data-efficient strategy that combines preprocessing to reduce frame redundancy with self-supervised diffusion learning to extract generalizable ultrasound feature representations. This approach effectively mitigated overfitting and achieved superior performance compared with benchmark transfer learning and self-supervised learning models under limited samples condition. Importantly, this is the first demonstration of a framework that integrates patient demographic, clinical, and full ultrasound video information for myofascial pain classification. While external validation will require larger multi-center datasets, our results provide preliminary but compelling evidence that deep learning can be adapted for small, prospectively collected cohorts as a proof-of-concept for testing early-stage clinical ideas. These findings highlight the promise of diffusion-based self-supervised learning in enhancing ultrasound video analysis, lay the groundwork for developing video-based biomarkers for reliable MTrP detection, chart a path toward scalable tools for myofascial pain diagnosis and management, and bridge the gap between early feasibility assessment and large-scale clinical validation.

## Declaration of generative AI and AI-assisted technologies in the writing process

During the preparation of this work the author(s) used ChatGPT in order to enhance manuscript readability. After using this tool/service, the author(s) reviewed and edited the content as needed and take(s) full responsibility for the content of the published article.

## Code and Data Availability

The data and code in this study are available upon request from the corresponding author.

## Data Availability

All data produced in the present study are available upon reasonable request to the corresponding authors

## Acknowledgments

No additional individuals or organizations contributed to this work. This work was not supported by external or internal funding and was not preregistered.

## Authors’ Contributions

**Hao-En Lu:** Writing – original draft, Writing – review & editing, Conceptualization, Investigation, Methodology, Visualization, Formal analysis. **David Koivisto:** Data curation, Resources. **Yuyang Lou**: Writing – review & editing. **Zixue Zeng:** Writing – review & editing. **Jing Wang:** Writing – review & editing. **Tong Yu:** Writing – review & editing. **Xin Meng**: Writing – review & editing. **Cameron Nowikow:** Writing – review & editing. **Dinesh Kumbhare:** Data curation, Writing – review & editing, Resources. **Richard Wilson** Writing – review & editing. **Jiantao Pu:** Writing – original draft, Writing – review & editing, Supervision, Funding acquisition, Conceptualization, Resources, Visualization, Project Administration.

## References

Arnab A, Dehghani M, Heigold G, Sun C, Lučić M, Schmid C. Vivit: A video vision transformer. Proceedings of the IEEE/CVF international conference on computer vision 2021: 6836–6846.

Behr M, Noseworthy M, Kumbhare D. Feasibility of a Support Vector Machine Classifier for Myofascial Pain Syndrome: Diagnostic Case-Control Study. J Ultrasound Med 2019;38: 2119–2132.

Chen T, Kornblith S, Norouzi M, Hinton G. A Simple Framework for Contrastive Learning of Visual Representations. Proceedings of the 37th International Conference on Machine Learning 2020;119: 1597–1607.

Dua A and Chang K-V. Myofascial pain syndrome. In: StatPearls [Internet]. StatPearls publishing; 2025.

Ericsson L, Gouk H, Loy CC, Hospedales TM. Self-Supervised Representation Learning: Introduction, advances, and challenges. IEEE Signal Processing Magazine 2022;39: 42–62.

Fuest M, Ma P, Gui M, Schusterbauer J, Hu VT, Ommer B. Diffusion models and representation learning: A survey. arXiv preprint arXiv:240700783 2024.

Gromakovskis V. Exploring fascia in myofascial pain syndrome: an integrative model of mechanisms. Frontiers in Pain Research 2025;6: 1712242.

Han A, Huang W, Cao Y, Zou D. On the feature learning in diffusion models. arXiv preprint arXiv:241201021 2024.

Hara K, Kataoka H, Satoh Y. Learning spatio-temporal features with 3d residual networks for action recognition. Proceedings of the IEEE international conference on computer vision workshops 2017: 3154–3160.

Harden RN. Muscle Pain Syndromes. American Journal of Physical Medicine & Rehabilitation 2007;86: S47–S58.

He K, Chen X, Xie S, Li Y, Dollár P, Girshick R. Masked autoencoders are scalable vision learners. Proceedings of the IEEE/CVF conference on computer vision and pattern recognition 2022: 16000–16009.

Ho J, Jain A, Abbeel P. Denoising diffusion probabilistic models. Advances in neural information processing systems 2020;33: 6840–6851.

Jain S, Li X, Xu M. Knowledge transfer from macro-world to micro-world: enhancing 3D Cryo-ET classification through fine-tuning video-based deep models. Bioinformatics 2024;40: btae368.

Kay W, Carreira J, Simonyan K, Zhang B, Hillier C, Vijayanarasimhan S, Viola F, Green T, Back T, Natsev P. The kinetics human action video dataset. arXiv preprint arXiv:170506950 2017.

Kim HE, Cosa-Linan A, Santhanam N, Jannesari M, Maros ME, Ganslandt T. Transfer learning for medical image classification: a literature review. BMC medical imaging 2022;22: 69.

Koh RG, Dilek B, Ye G, Selver A, Kumbhare D. Myofascial Trigger Point Identification in B-Mode Ultrasound: Texture Analysis Versus a Convolutional Neural Network Approach. Ultrasound in Medicine & Biology 2023;49: 2273–2282.

Krishnan R, Rajpurkar P, Topol EJ. Self-supervised learning in medicine and healthcare. Nature Biomedical Engineering 2022;6: 1346–1352.

Lucas N, Macaskill P, Irwig L, Moran R, Bogduk N. Reliability of physical examination for diagnosis of myofascial trigger points: a systematic review of the literature. Clin J Pain 2009;25: 80–89.

Mukhopadhyay S, Gwilliam M, Yamaguchi Y, Agarwal V, Padmanabhan N, Swaminathan A, Zhou T, Ohya J, Shrivastava A. Do text-free diffusion models learn discriminative visual representations? European Conference on Computer Vision 2024: 253–272.

Rathbone AT, Grosman-Rimon L, Kumbhare DA. Interrater agreement of manual palpation for identification of myofascial trigger points: a systematic review and meta-analysis. The Clinical journal of pain 2017;33: 715–729.

Sampat MP, Wang Z, Gupta S, Bovik AC, Markey MK. Complex wavelet structural similarity: A new image similarity index. IEEE transactions on image processing 2009;18: 2385–2401.

Satopaa V, Albrecht J, Irwin D, Raghavan B. Finding a “kneedle” in a haystack: Detecting knee points in system behavior. 2011 31st international conference on distributed computing systems workshops 2011: 166–171.

Schisterman EF, Faraggi D, Reiser B, Hu J. Youden Index and the optimal threshold for markers with mass at zero. Statistics in medicine 2008;27: 297–315.

Selvaraju RR, Cogswell M, Das A, Vedantam R, Parikh D, Batra D.Grad-cam: Visual explanations from deep networks via gradient-based localization. Proceedings of the IEEE international conference on computer vision; 2017; 618–626.

Shea DE, Kulhare S, Millin R, Laverriere Z, Mehanian C, Delahunt CB, Banik D, Zheng X, Zhu M, Ji Y. Deep learning video classification of lung ultrasound features associated with pneumonia. Proceedings of the IEEE/CVF conference on computer vision and pattern recognition 2023: 3103–3112.

Shomal Zadeh F, Koh RG, Dilek B, Masani K, Kumbhare D. Identification of Myofascial Trigger Point Using the Combination of Texture Analysis in B-Mode Ultrasound with Machine Learning Classifiers. Sensors 2023;23: 9873.

Sikdar S, Shah JP, Gebreab T, Yen R-H, Gilliams E, Danoff J, Gerber LH. Novel applications of ultrasound technology to visualize and characterize myofascial trigger points and surrounding soft tissue. Archives of physical medicine and rehabilitation 2009;90: 1829–1838.

Tong Z, Song Y, Wang J, Wang L. Videomae: Masked autoencoders are data-efficient learners for self-supervised video pre-training. Advances in neural information processing systems 2022;35: 10078–10093.

Xiang W, Yang H, Huang D, Wang Y. Denoising Diffusion Autoencoders are Unified Self-supervised Learners. 2023 IEEE/CVF International Conference on Computer Vision (ICCV) 2023: 15756–15766.

Zu W, Xie S, Chen H, Ma L. Pre-trained Models Succeed in Medical Imaging with Representation Similarity Degradation. arXiv preprint arXiv:250307958 2025.

